# Evaluating Research Ethics Committees Using a Newly Developed Assessment Tool: Experience from Sri Lanka

**DOI:** 10.64898/2026.01.14.26344139

**Authors:** CA Wanigatunge, P Ranatunge, PHGJ Pushpakumara, W Subasinghe, HH Peiris, CL Dandeniya, S Prathapan

## Abstract

**Introduction:** Effective research ethics committees (RECs) are essential for safeguarding research participants and maintaining public trust in health research. The economic crisis in Sri Lanka made external accreditation financially difficult, and a locally led, affordable approach was required to strengthen ethics oversight and quality assurance.

**Methods:** The Forum for Research Ethics Committees in Sri Lanka developed a locally led framework and digital assessment tool to evaluate REC practices. Indicators were selected through a modified Delphi process, based on World Health Organization’s “Guidance for Evaluating Ethical Review Practices” and the “Benchmarking Tool for Ethics Oversight of Health-related Research”. Indicators assessed REC structure, resources, procedures, transparency, performance monitoring, and completeness of the ethics review process. A secure digital tool was developed and piloted through on-site surveys of two RECs representing different institutional maturity levels and medical systems and was refined for local applicability.

**Discussion/ Findings:** The framework enabled systematic assessment of both institutional readiness and actual ethics review practices. Incorporating document review and direct observation allowed identification of strengths and gaps not captured by benchmarking indicators alone. The process was feasible, resource-appropriate, and supported national capacity building through training of surveyors. The paperless digital approach improved standardisation, traceability and feasibility at low cost.

**Conclusion:** The FRECSL mechanism provides a transparent, low-cost approach to quality assurance of RECs in resource-limited settings. By combining readiness assessment with evaluation of review practice, it offers a balanced model aligned with international standards and may be adapted by other countries seeking to strengthen ethical oversight of health-related research.

## Introduction

Research ethics committees (RECs) - also referred to as Ethics Review Committees (ERCs) or Institutional Review Boards (IRBs) - are integral to the research governance landscape. They are mandated to safeguard the rights, safety, and well-being of research participants and are expected to function independently of the institutions to which they are affiliated. In accordance with international best practice, in Sri Lanka too, ethical approval is routinely sought before the commencement of human research. To maintain public trust and meet the expectations of regulators and funding agencies, RECs must operate to standards that ensure rigorous and transparent ethical review of research protocols.

In Sri Lanka, RECs are affiliated with both universities and hospitals. They operate according to the guidelines set by the World Medical Association in their Declaration of Helsinki (1) and the operational guidelines provided by the Forum for Research Ethics Committees in Sri Lanka (FRECSL) (2). FRECSL is an independent forum convened under the Sri Lanka Medical Association (SLMA) to foster improved understanding and implementation of ethics review of biomedical research in Sri Lanka.

Quality assurance is a crucial public duty of RECs and involves continuing education of committee members, internal monitoring, and periodic external reviews carried out by independent organisations. External review mechanisms are intended to provide constructive guidance to strengthen REC performance and to offer reassurance to researchers, institutions, and funders that ethical review processes meet accepted standards. However, there is no universally accepted method to evaluate whether RECs operate effectively and within acceptable standards. While institutional readiness and capacity can be evaluated, assessing the quality and completeness of ethics review in practice remains methodologically challenging.

In the absence of a national accreditation or regulatory system, several Sri Lankan RECs previously sought external evaluations through the Strategic Initiative for Developing Capacity in Ethical Review (SIDCER) programme, facilitated by the Forum for Ethics Review Committees in Asia and the Western Pacific (FERCAP) (3). These assessments, based on WHO-TDR operational guidelines (4) were supported by FRECSL and provided evidence-based recommendations to RECs to improve their capacity and standards. The SIDCER reviews also enabled members of Sri Lankan RECS to serve as reviewers for SIDCER’s evaluations of regional RECs, thereby enhancing the national capacity.

Sri Lankan RECs joined the SIDCER reviews in 2009 and continued until the COVID pandemic in 2019, and nine RECs were assessed at regular intervals. Eight of these were affiliated with medical faculties or universities, and one with the country’s apex professional organisation. Although not a regulatory requirement and despite the high cost involved, RECs in SL voluntarily participated in these surveys to enhance their capacity and the quality of ethics review

Following Sri Lanka’s economic crisis in 2021, the continuation of paid external accreditation became unsustainable even for the RECs that had previously undergone SIDCER review. In response, FRECSL was tasked with developing a locally led, resource-appropriate mechanism to evaluate REC performance. This paper describes the development, piloting, and implementation of a national framework and assessment tool to survey and evaluate the practices of RECs in Sri Lanka, with potential applicability to other resource-limited settings.

## Method

### Framework development and indicator selection

FRECSL appointed a multidisciplinary subcommittee to develop this framework. The subcommittee reviewed the WHO-TDR Surveying and Evaluating Ethical Review Practices: Operational Guidelines (2002) (4) and the WHO Benchmarking Ethics Oversight of Health-Related Research Involving Human Participants tool (2023) (5). Both documents shared common assessment domains, but the latter also classified the indicators according to levels of implementation (fully, partially, or not implemented). The Declaration of Helsinki (1) and the FRECSL operational guidelines (2) were used to define minimum standards.

A modified Delphi technique was used to determine which domains and indicators should be included. The expert panel comprised health professionals with extensive experience in research ethics across multiple medical disciplines. In the first round, panel members assessed whether each domain was necessary for inclusion. There was unanimous agreement to exclude the first and the last categories in the WHO tool for benchmarking (5) – i.e. categories 1 and 7, which were related to the legal and regulatory framework and the responsible research institution, respectively, as these were outside the mandate of FRECSL and therefore of the surveys.

Subsequent Delphi rounds focused on the selection and refinement of indicators within the remaining domains, using a binary yes/no scale. Indicators lacking consensus were revised based on panel feedback and re-circulated. Indicators were excluded if agreement did not reach 75% after revision.

### Assessment categories and operationalisation

Five categories were selected from section 2 of the WHO benchmarking tool – indicators 2-6 (6). In addition, the subcommittee included an assessment of the completeness of the ethics review process, drawing on prior SIDCER experience and FRECSL guidance. This component aimed to evaluate review practice rather than organisational readiness alone. Table 1 summarises the categories identified for evaluation, and the documents perused and observations made to evaluate these categories.

**Table 1:** Categories to be evaluated when surveying a REC.

| **Categories to be evaluated \*** | **No of indicators** | **Documents perused and observations made** |
| --- | --- | --- |
| REC structure and composition<br>(4,6) | 5 | The self-assessment form submitted by REC, SOPs, membership files, organogram of REC, types of protocols reviewed by REC, evidence for external reviews when expertise was not available |
| REC resources (4,6) | 5 | REC staff training records, interview of REC staff, visiting the REC office |
| REC procedures (4,6) | 7 | Website, SOPs, protocol evaluation |
|  |  | forms, communication logs, REC's document management system |
| Completeness of review process (4) | 2 | Protocol file review, observation of REC meeting |
| Mechanisms to promote REC's transparency and accountability (4,6) | 6 | Website (if available), SOPs, communication logs/files |
| Mechanisms for RECs to monitor their performance (6) | 3 | SOPs, communication logs/files, reports of internal audits |

\* The relevant WHO guidance is given in parentheses, REC: research ethics committee, SOPs: standard operating procedures of the REC surveyed

To operationalise the selected criteria and ensure consistency across surveys, a customised digital data collection tool was developed. The digital instrument included structured data-validation rules, conditional logic, and embedded quality checks to support uniform documentation of evidence, observations, and interview findings, thereby reducing inter-reviewer variability. Automated summary views were integrated to enable real-time visualisation of compliance patterns across the eight identified criteria categories, enhancing transparency and supporting robust audit trails. By adopting a fully paperless workflow, the tool reduced the environmental impact of the survey while improving standardisation, reproducibility, and traceability of REC evaluations in line with recognised international frameworks (4,5). The data collection tool was password-protected with access restricted to authorised survey team members. Technical and procedural safeguards were implemented to maintain confidentiality and data integrity.

### Indicator scoring

Each indicator was categorised as not implemented, partially implemented, or fully implemented, in line with WHO benchmarking guidance (5). Although some practices—such as public disclosure of funding sources and formal internal audits—are not routinely implemented by RECs in Sri Lanka, these indicators were retained as they represent internationally recognised good practice.

### Pilot testing

The assessment tool was piloted on two RECs that had requested to be surveyed. One was a well-established REC affiliated with the allopathic medical system that had previously undergone two SIDCER reviews and reviewed biomedical research protocols exclusively within the allopathic system. The second was a newer REC, established four years earlier, affiliated with an institution of Ayurvedic and traditional medicine and responsible for reviewing research in traditional and alternative medical systems. These two contexts reflect the main forms of health-related research conducted in Sri Lanka. Minor modifications were made to the indicators and data-collection tool following the pilot surveys to improve clarity, relevance, and suitability for the local context.

### Survey procedure

The survey process was modelled on the SIDCER surveys with which Sri Lankan RECs were familiar (4). Each survey team comprised three surveyors and a FRECSL coordinator, supported by nine assistant surveyors. Surveyors were selected from individuals with prior experience in SIDCER reviews, while assistant surveyors included former trainees and new participants. All survey team members received initial training in the new survey tool and methodology, and the surveys were conducted on-site over three days.

The survey commenced with an opening meeting during which the FRECSL coordinator outlined the objectives and procedures, and the REC chair presented the committee’s mandate, scope, and functions. The survey teams assessed REC operations through review of standard operating procedures and other documentation, site visits to REC offices, interviews with REC staff, and observation of a full board meeting.

At the end of each day, survey teams met in closed session to review findings, discuss evidence, and reach consensus on indicator scoring. The FRECSL coordinator supervised the process and provided ongoing guidance, while also supporting capacity building among assistant surveyors.

### Reporting and follow-up

Preliminary findings were presented to REC members and institutional representatives at the conclusion of each survey. This facilitated open discussion and provided RECs with an opportunity to clarify practices or respond to observations. A detailed written report, supported by documentary and observational evidence, was subsequently provided to each REC. Reports identified good practices and areas requiring improvement, and RECs were requested to submit an action plan outlining corrective measures.

The action plans were reviewed by the FRECSL subcommittee, and final determinations were communicated to the RECs. This process emphasised transparency, dialogue, and continuous quality improvement rather than punitive assessment.

## Discussion

This paper describes the development and implementation of the first locally led system for evaluating the performance of RECs in Sri Lanka. The FRECSL mechanism was designed to assess both organisational readiness and the conduct of ethics review in practice, using explicit, transparent criteria. Grading indicators (5) according to levels of implementation enabled RECs to identify gaps and prioritise improvements while providing assurance to funders, institutions, and regulators that ethical review processes align with accepted standards.

The WHO benchmarking tool (5) proved useful for assessing the preparedness of a REC for ethics review. However, as scoring is largely based on the existence of structures and procedures, it was insufficient to capture how the actual ethics review process operates and may provide a misleading estimate of the REC’s capabilities. By incorporating elements from the WHO-TDR guidelines (4) and prior SIDCER experience, the FRECSL framework enabled assessment of the completeness and quality of ethical review through document review and direct observation. This combined approach allowed a more nuanced and accurate appraisal of REC performance.

Ensuring independence of review was a key challenge given the limited national pool of experts. Although the inclusion of international surveyors could have enhanced perceived impartiality, financial constraints precluded this option. Potential bias was mitigated through transparent selection of survey teams, advance disclosure of these teams to RECs, use of objective indicators, and evidence-based reporting. RECs were also afforded opportunities to challenge findings, reinforcing procedural fairness and credibility.

Capacity building was embedded within the process through the training of assistant surveyors, thereby expanding national expertise and supporting sustainability. In the absence of a statutory accreditation framework, FRECSL surveys remain voluntary but provide an important interim mechanism for quality assurance. It is hoped that Sri Lanka will develop a formal mechanism to accredit RECs, which will mandate all RECs to undergo a review/survey process. Establishing the legal framework for this needs to be identified as a priority in the field of research ethics by policymakers.

## Conclusion

The FRECSL evaluation framework offers a feasible, low-cost approach to quality assurance of research ethics committees in resource-limited settings. By combining assessment of institutional readiness with evaluation of actual review practices, the mechanism provides a balanced and objective appraisal aligned with international standards. Although the absence of international reviewers is a limitation, the use of explicit criteria and transparent procedures supports independence and credibility. This model may be readily adapted by other countries seeking to strengthen ethical oversight of health-related research.

## Data Availability

All data produced in the present study are available upon reasonable request to the authors

## Author contributions

All authors (CAW, PR, PHGJP, WS, HHP, CLD, SP) conceptualised the survey, identified the indicators and developed the survey plan.

PR and PHGJP developed the digital tool.

SP, PR, and WS were surveyors of the pilot surveys, PHGJP and CAW were FRECSL coordinators for the pilot surveys.

CW, SP, PR and WS developed the initial draft.

All authors reviewed and approved the final manuscript.

## Ethical consideration

The activity was identified as a priority by the Management Committee of FRECSL as per its mandate. As it was not research and there was no involvement of individuals, ethics approval was not obtained for the development of the tool and its piloting.

## Acknowledgements

The authors wish to acknowledge:

- the Sri Lanka Medical Association and the Bandaranaike Memorial Research Institute, Sri Lanka and their Research Ethics Committees for supporting the piloting of the tools
- the surveyors and the survey teams for their support in helping to fine tune to survey tool and methodology

